# Validity of AI-based long-term cough monitoring for clinical use

**DOI:** 10.1101/2025.07.25.25332190

**Authors:** Mitja Alge, Laura Kuett, Alexander Duschau-Wicke, Michael Häberli, Alyn Morice, Surinder Birring, David D. Elkayam

## Abstract

Chronic cough is a debilitating condition that affects quality of life and often signals underlying respiratory disease. Clinical studies currently rely on audio recordings reviewed by human analysists to objectively quantify cough frequency, but this process is time-consuming and limits long-term monitoring. Emerging automated cough measurement tools enable multi-day measurements and therefore offer a more accurate assessment of a patient’s actual cough burden. At the same time, they are expected to show varying performance across patients. The tradeoff between algorithmic accuracy and the number of measurement days required to obtain a valid estimate of an individual’s objective cough burden has not previously been explored. In this study, we aimed to validate the performance of our proprietary cough detection algorithm, part of an automated cough monitoring system, in patients with chronic cough. We also investigated how algorithm performance and daily variability in cough frequency affect the optimal number of monitoring days. The algorithm was evaluated under real-life conditions in 51 patients with chronic cough, achieving a median sensitivity of 0.93 and a median precision of 0.94. By using a confidence score to identify unreliable data and applying bootstrapping simulations to model variability, we found that the largest gains in measuring true cough burden were achieved with three days of monitoring, with diminishing returns beyond seven days. In conclusion, because cough frequency fluctuates daily, automated cough counting tools can enhance human-annotated data by enabling multi-day monitoring to more accurately capture true cough burden.

## Introduction

Chronic cough has been defined as a cough that lasts for eight weeks or longer and is associated with many underlying respiratory diseases, including chronic obstructive pulmonary disease, asthma, and idiopathic pulmonary fibrosis, with an estimated global prevalence of approximately 5-10% (Song et al. 2015). Patients with chronic cough may cough from hundreds to thousands of times per day (Hsu et al. 1994). Objective cough counts have become a primary endpoint in clinical trials of anti-tussive therapies (Dicpinigaitis et al. 2023; McGarvey et al. 2023; Abdulqawi et al. 2015) and have been suggested as a useful marker to better understand the aetiologies and treatment efficacy in the clinics (Morice et al. 2007; 2020). There has been an emergence of several automated cough measurement tools which enables the capture of cough counts over extended periods (Birring et al. 2008; Gross et al. 2019; Kuhn et al. 2023; Chaccour et al. 2025). The measurement of cough over these extended periods has revealed that there is a high day to day variability in cough counts (Morice et al. 2025; Chung et al. 2024). Thus, to more accurately estimate the true cough burden suffered by an individual we suggest that it should be averaged across multiple days. The measurement of cough over longer periods is only feasible by automated tools due to the time and effort required to obtain accurate data by human listeners manually identifying cough events (Mines et al. 2019). It is expected that automated tools will have varying performance across patients, yet because of their ability to capture much more data they will enable a more accurate capture of the actual cough burden. The tradeoff between accuracy and the multi-day measurement to achieve a valid readout of individuals’ objective cough has not been examined before. Therefore, we aimed to evaluate the performance of our cough detection algorithm as part of our proprietary cough monitoring system in patients with chronic cough, and to examine how the performance and daily cough variability informing us about the optimal number of measurement days that are sufficient to capture cough burden.

## Results

### Cough algorithm’s clinical validation setup

We developed a Deep Neural Network-based ensemble model to detect cough explosions from audio data. This model is a key part of the SIVA System. The SIVA System comprises a wearable device that collects audio data, which is uploaded to a cloud-based backend where the cough algorithm processes the data, providing timestamps for each detected cough explosion. The model was trained on publicly available and proprietary data, including data specifically collected with the SIVA wearable device. The entire system was developed as part of a quality-controlled process aligned with ISO 13485, including documentation, testing, and validation steps required for medical device certification.

To evaluate the performance of the cough detection algorithm as part of the SIVA System, we conducted a clinical validation study in which patients with chronic cough used the SIVA System under real-life conditions. In this study, patients were requested to wear the proprietary SIVA Wearable cough monitor device around their neck at the start of visit 1 and place it on a wireless charger at their bedside during nighttime, where the device continued to collect cough data. The observation period comprised of 14 consecutive days, however, only the first 24h were used for the algorithm validation evaluation. A total of 52 patients were recruited for the study; audio data from one patient was not collected due to a technical failure of the wearable device. Out of the 51 patients who completed the study (80% women, median age 66), 7 patients had asthma, in addition to a diagnosis of chronic cough (Table 1). The algorithm performance was evaluated by comparing cough explosion timestamps annotated by human labelers to the cough algorithm output.

**Table 1.** Study population data.

| Baseline Characteristics | Value |
| --- | --- |
| Number of patients | 51 |
| Median age (min-max) | 66 (21-84) |
| Female, number (%) | 40 (80%, 1 unknown) |
| Diagnosis or suspicion of chronic cough, number (%) | 51 (100%) |
| Diagnosis of asthma, number (%) | 7 (14%) |
| Median Cough Severity VAS 2 weeks prior to the study in mm (min-max) | 60.6 (8.4- 98.4) |
| Median Leicester Cough Questionnaire total score 2 weeks prior to the study, number out of 21 (min-max) | 13.5 (5.5-20.2) |

### Confidence score flags patients with low algorithm reliability

An inherent risk of machine-learning models is that they may not maintain their expected performance when confronted with input data that substantially deviates from their training data. Despite the extensive body of training data used in the development of our cough detection algorithm, prior work has indicated that some individuals have cough characteristics that strongly deviate from commonly observed cough patterns. To account for such exceptions and increase the overall robustness of our cough detection algorithm, we have applied an approach commonly used in machine learning: using an ensemble model consisting of differently trained individual models and evaluating the degree of agreement of the individual models in the ensemble to rate the confidence of the overall decision of the algorithm. The resulting confidence score is intended to represent how well the cough detection algorithm performed on a given day for a given patient.

First, we evaluated how well the confidence score represents the cough detection performance. The final model performance score (F1 score, the harmonic mean of sensitivity and positive predictive value) correlated strongly with the confidence score across patients (Pearson R = 0.74). Second, we explored the reasons why the six patients were flagged as having a low or medium confidence score in the context of the algorithm’s performance (Figure 1, P1-P6). By examining the audio data captured during the first 24 hours, we could identify common causes why the cough detection algorithm was uncertain about its predictions. Despite the adequate performance score, Patient 1 had a low confidence score due to the presence of very loud background noise from a music concert, creating a challenging scenario for the algorithm. With varying performance levels, patients 2, 3, and 5 all exhibited very frequent, strong, and explosive throat-clearing sounds that were challenging for the algorithm to discriminate from coughs. Patient 3 experienced especially low performance due to the similarity of the throat-clearing sounds to coughing. Patients 4 and 6 had many coughs in the background that did not belong to the patient. However, the algorithm was trained to detect all cough explosions regardless of the subject, which led to a difference in performance. For patient 4, those coughs were handled as cough explosions, but for patient 6, the quiet background coughs were not detected by the algorithm.

**Figure 1.**
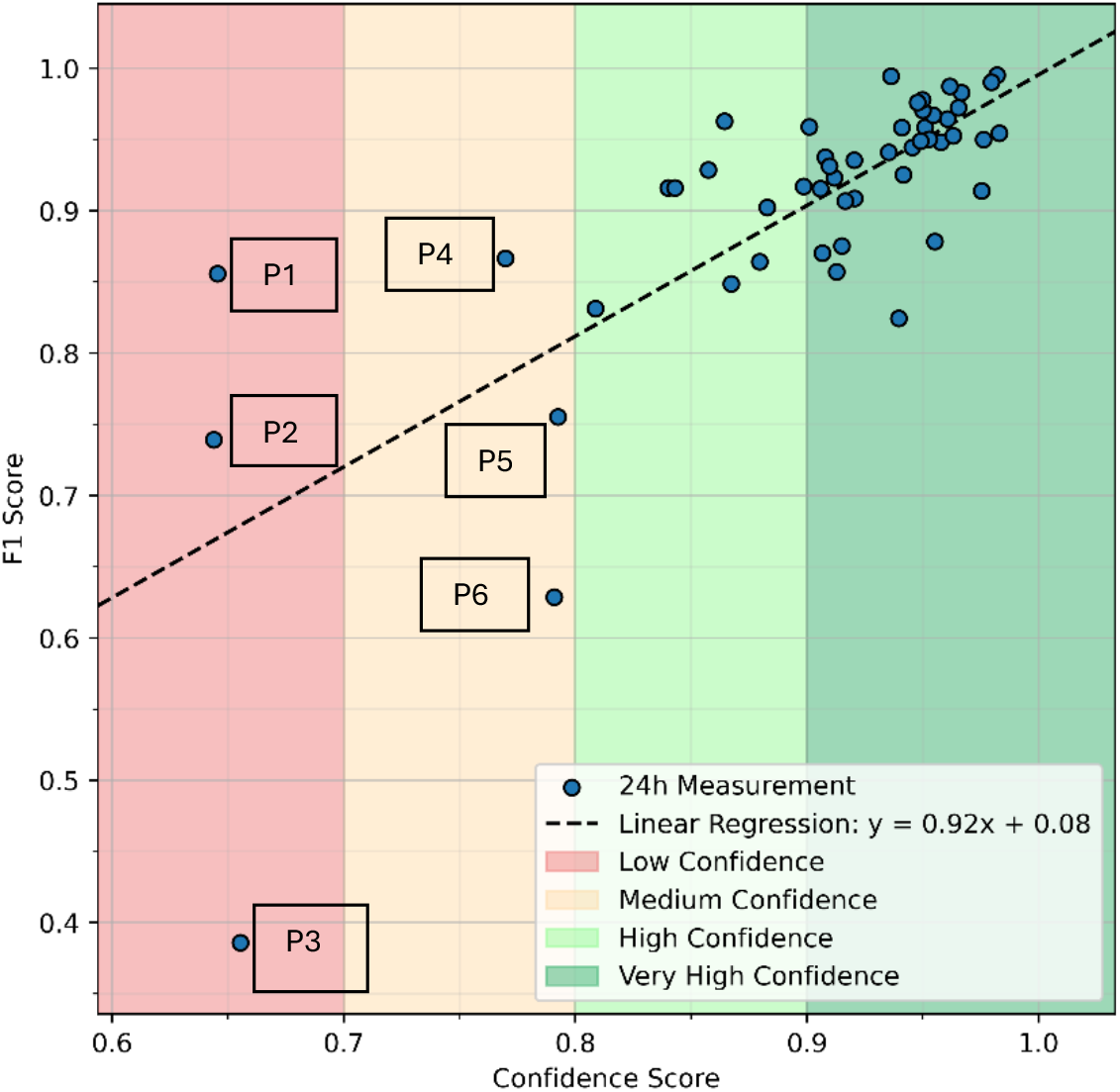
Confidence score correlates with the algorithm’s performance. Correlation plot showing F1 Score vs Confidence Score (Pearson R = 0.74). Each dot represents the performance (F1 score) of the cough detection algorithm in detecting each cough explosion for an individual patient (n = 51). The confidence score is calculated as the average uncertainty score across the ensemble model for the 24h period for each patient. Patients with low and medium confidence scores are marked in the boxes.

### Performance of the cough detection algorithm

First, as the cough frequency (total number of coughs divided by the recording period, typically 24 hours) has been used as a standard metric in clinical trials, we examined the agreement for capturing accurate cough frequency between the algorithm output and human labelers. To achieve this, we calculated the concordance correlation coefficient (CCC) between the algorithm-based and the human-labeled cough frequencies. The CCC value was 0.988, indicating that the cough detection algorithm accurately captured the cough frequency in this study population (Figure 2).

**Figure 2.**
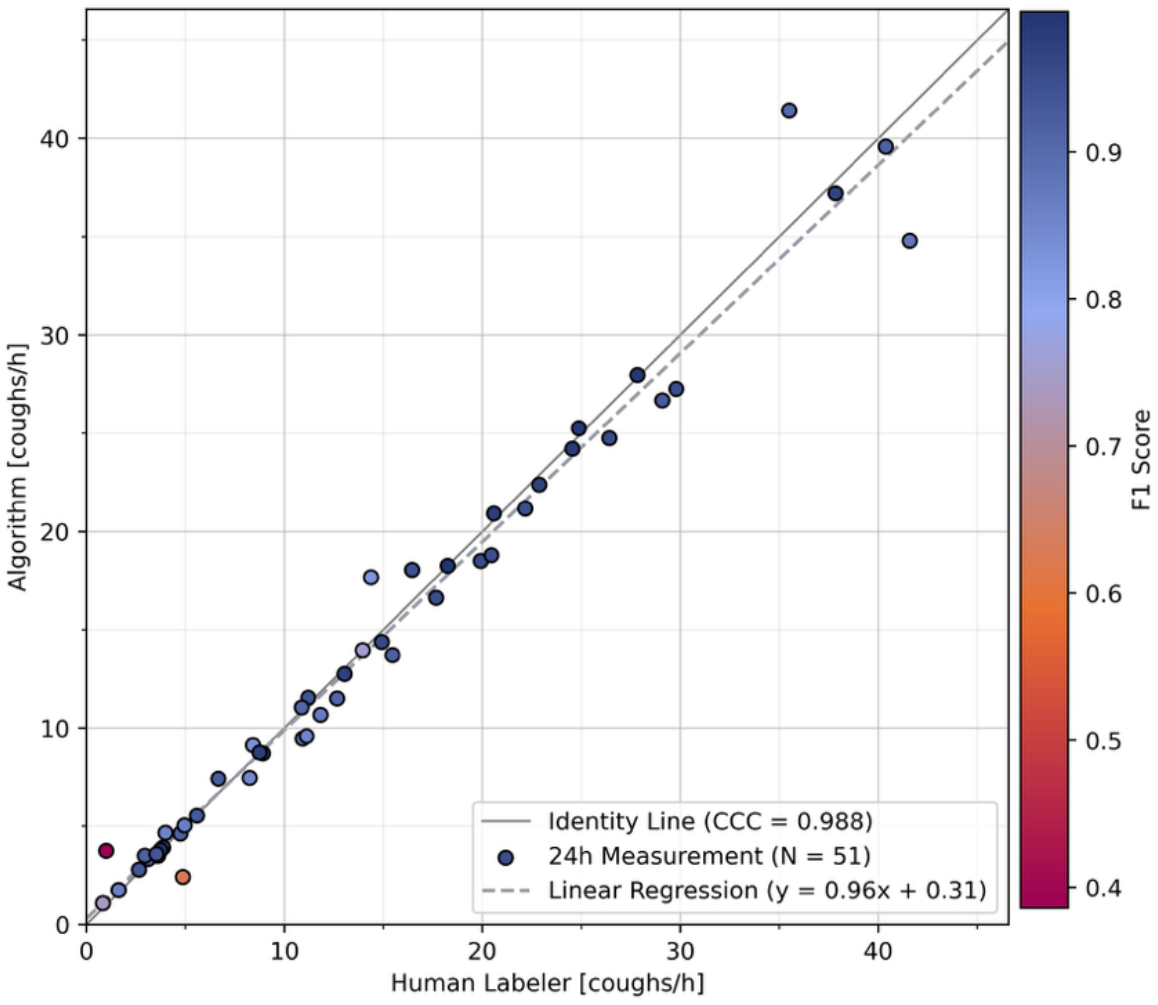
Number of coughs detected by the algorithm vs human labellers. A concordance correlation coefficient (CCC) plot comparing algorithm-detected coughs with human labelers. Each dot represents the cough frequency of cough explosions for each patient, as detected by the algorithm versus counted by human labelers (n = 51). The dots are coloured by the performance metric (F1 score). CCC= 0.988

To further examine the algorithm’s performance across low and high cough frequencies, we plotted the cough frequency as counted by human labelers against the F1 score (Figure 3). This indicated that the algorithm’s performance was consistent across different cough frequencies, except for patients with low confidence scores.

**Figure 3.**
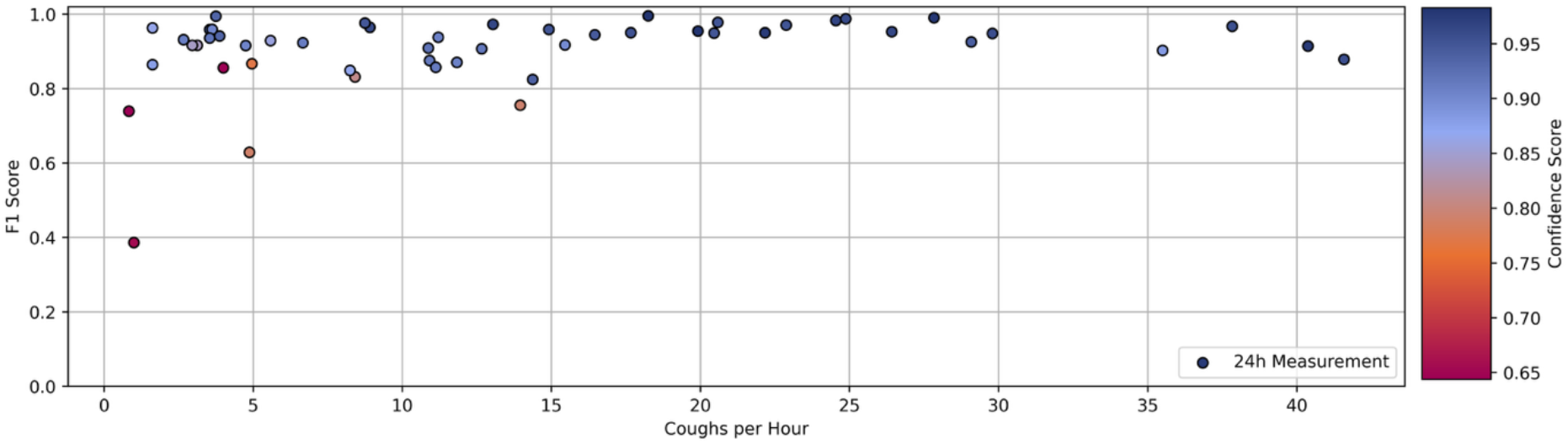
The algorithm’s performance for different cough frequencies. Each dot represents the cough frequency of cough explosions for each patient (n = 51), as counted by human labelers on the x-axis, and the algorithm’s performance metrics (F1 score) on the y-axis. The dots are coloured by the confidence score.

**Figure 4.**
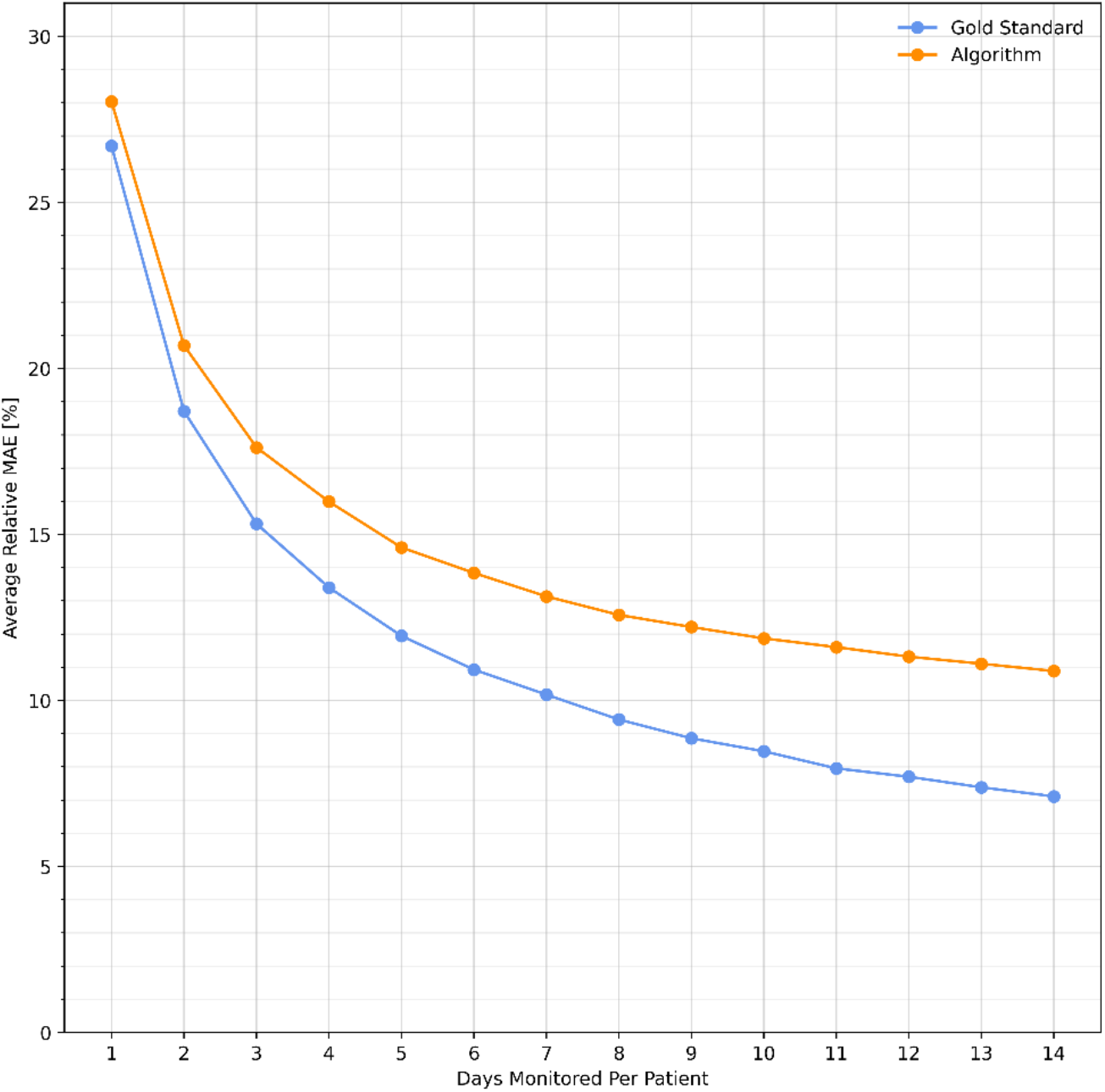
Impact of monitoring duration on cough burden estimation accuracy. Number of days monitored on × axis, average relative mean absolute error (MAE) on y axis. The relative mean absolute error (MAE) was computed for each duration (1-14 days), applying a measurement error model that reflects algorithm performance (orange line) and for a hypothetical gold standard with perfect cough detection (blue line). The error is calculated in comparison to the hypothetical true cough burden, derived as the average of the 14-day cough frequency for each patient, as measured with the SIVA system in the clinical validation study.

Second, as the algorithm detects each cough explosion and outputs a timestamp for this event at millisecond resolution, we evaluated how well the algorithm detects cough explosions in each patient. We calculated sensitivity (recall), positive predictive value (precision), and F1 score for all patients. Additionally, we examined the performance of the algorithm when patients were grouped by at least medium, high, or very high confidence score levels (Table 2). Across all patients, the algorithm achieved a median sensitivity of 0.93 and a median precision of 0.94. Our analysis showed that including only patients with at least a medium confidence score improves the algorithm’s performance for both sensitivity and precision showing the usefulness of detecting outlier data using the confidence score, but increasingly marginal gains are achieved when only including patients with high or very high confidence score levels.

**Table 2.**
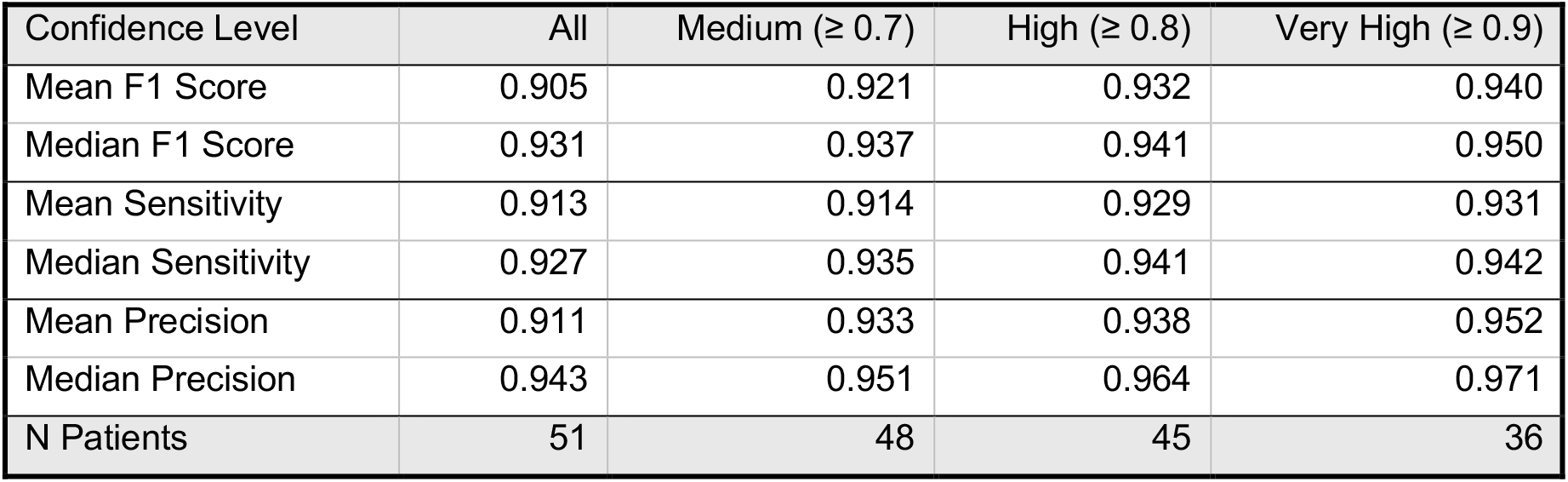
The cough detection algorithm performance metrics.

### Quantifying the impact of monitoring time and algorithm performance on capturing true cough burden

Clinical trials for chronic cough drugs have, so far, used human-labeled data that is considered close to 100% accurate; however, this data is limited to a single 24-hour period due to the time-consuming efforts required for cough counting. Automated tools would enable multi-day measurements with less than 100% accuracy, yet capture the daily cough variability, thus providing a more stable measure if averaged across days. Therefore, we examined how monitoring length influences the accuracy of estimated average cough frequency (“cough burden”) while explicitly accounting for day-to-day variability and algorithm measurement error. Using a bootstrap-based simulation with real patient data (n = 48) from 14-days of cough monitoring and after excluding patients with a low confidence score, we sampled up to 14 days of patient-level data and calculated the normalized deviations of the averages from each patient’s 14-day mean (“true cough burden”) to yield an Average Relative MAE curve. We compared the Average Relative MAE curves for the algorithm (including the measurement error resulting from less-than-perfect algorithm performance) and for the hypothetical gold standard (i.e., with assumed perfect cough detection performance). Consequently, the analysis separates the contribution of algorithm performance from the contribution of natural biological variability to measurement error, enabling the understanding of the effect of performance on the number of optimal monitoring days required. Our analysis shows that a single-day ‘perfect’ measurement would be off from the true cough burden by about 27%, whereas incorporating a potential algorithm performance error would only increase the error to 28%. Additionally, we found that the largest improvements in measuring true cough burden were achieved with three days of automated monitoring, resulting in an error reduction from 28% to approximately 18%. The error rate then decreased to 13% by day 7, with diminishing returns beyond that point. A 7-day measurement period with an automated tool results in approximately half the measurement error compared to a 1-day human labeled tool (error is reduced from ~27% to 13%). The analysis further highlights that, due to natural variability in cough frequency, a 14-day automated measure would still have an average error of 11% compared to the gold standard with an error of 7%.

## Discussion

In this study, the cough detection algorithm of a fully automated cough monitoring system was evaluated under real-life conditions in 51 patients with chronic cough, achieving a median sensitivity of 0.93 and a median precision of 0.94. By using a confidence score to identify unreliable data and applying bootstrapping simulations to model variability, we found that the largest improvements for measuring true cough burden were achieved with three days of monitoring, with diminishing returns beyond seven days. This contributes to understanding how to effectively incorporate automated cough-counting tools into clinical trials and healthcare settings to accurately measure the true cough burden.

Our study has several limitations. First, the validation of our cough detection algorithm relies on audio data collected using a proprietary wearable device. Ideally, an external reference device would ‘have been used during the first 24 hours to confirm whether all coughs were captured. However, internal bench testing, the microphone specifications, and the audio data itself indicate that even very quiet coughs were reliably detected by the device. Additionally, our validation dataset included only patients with chronic cough with or without a diagnosis of asthma. The algorithm should be further validated in other respiratory conditions where cough is a common symptom, such as idiopathic pulmonary fibrosis (IPF), chronic obstructive pulmonary disease (COPD), and bronchiectasis. Nonetheless, the algorithm’s training data included patients with these conditions, and we do not expect significant inherent differences in cough acoustics across this population aside from general variability in algorithm performance, as reflected in the confidence score analysis. Regarding the confidence score, future work could refine the heuristics used to define low, medium, and high-performance categories and assess the score’s effectiveness in identifying outlier patients, with the goal of improving performance flagging. Despite using simple heuristics, we were able to identify patients with high background noise, frequent throat-clearing sounds resembling coughs, and substantial background coughing from others. Finally, our bootstrapping simulation could be enhanced by including patients with long-term cough monitoring over several months to better characterize the true cough burden in patients with chronic cough.

In this study, we demonstrate that additional metrics such as the confidence score can support the integration of machine learning-based technologies into clinical trials and healthcare practice by enhancing the reliability of data collected through these methods. Our analysis also highlights the need to develop algorithms capable of detecting throat-clearing sounds and distinguishing coughs not originating from the target subject. While these sounds may affect algorithm performance, they could also offer valuable insights, for example, throat clearings may reflect throat irritation associated with a chronic cough (O’Hara et al. 2022). Furthermore, the confidence score could be extended into a more comprehensive uncertainty measure, such as a confidence interval for the reported cough frequency.

Finally, by using a confidence score to identify unreliable data and applying bootstrapping simulation to compare subsets of 1–14 monitoring days against each patient’s true cough burden and incorporating a measurement error model reflective of algorithm performance, we identified how estimation error decreases with longer monitoring periods. The largest improvements in measuring true cough burden were achieved with three days of monitoring, with diminishing returns beyond seven days. These results are expected to support determining the optimal number of monitoring days required to balance accuracy with practical feasibility in clinical trial and healthcare settings.

In conclusion, because cough frequency shows substantial natural variations from day to day, automated cough counting tools can enhance human-annotated data by enabling multi-day monitoring, thereby capturing the true cough burden more accurately.

## Methods

### Device description

The SIVA System comprises a wearable device, a cloud-based backend infrastructure, and a patient-facing smartphone application. The custom wearable is worn on the patient’s chest around their neck to capture accurate audio data. While the device is on a wireless charger, encrypted and recorded audio segments are automatically uploaded from the device using Wi-Fi to a secure cloud-based backend infrastructure. In the cloud infrastructure, a proprietary neural network-based cough detection algorithm processes the data and outputs the exact timestamps of each cough explosion with millisecond accuracy. The wearable device continuously records movement data with its built-in 3-axis accelerometer. This movement data is used to determine whether the wearable device is worn.

### Model input data

The cough detection algorithm was developed using a combination of publicly available and proprietary datasets. Public sources such as AudioSet (Gemmeke et al. 2017), Coughvid-V1 (Orlandic et al. 2023), and RADAR-V1 (Smith et al. 2021) were incorporated during the early stages of development. To supplement this, a custom dataset was collected from usability testers and volunteers, focusing on real-world audio, including coughing, laughing, sneezing, and throat clearing. These recordings were captured using a prototype device, a custom smartphone application, and the current device during usability testing. In addition, SIVA conducted a clinical validation study using a prototype device (Kuhn et al. 2023). Data from the first 24 hours of monitoring for each participant in that study were included in the algorithm development process.

### Model architecture and development

The cough detection algorithm is the subject of a patent application covering machine learning-based body action detection (Mohammadi et al. 2024). It is based on Deep Neural Networks optimized with the ADAM optimizer (learning rate = 0.001) and a binary cross-entropy loss function. The audio is split into 5-second windows with a 2.5-second shift for inference. The input features consist of a 2D mel spectrogram matrix (1250 × 64) and a raw wave input at 16 kHz (1x 80000).

The model employs a hybrid architecture, comprising a 2D deep convolutional network that processes the mel-spectrogram, resulting in a block size of 157 × 128, and a 1D deep convolutional network that processes the raw input, resulting in a block size of 157 × 64. Each convolutional layer is followed by batch normalization and ReLU activation. Max pooling layers are used to reduce the spatial dimensions of the features, and residual connections are incorporated in the 1D raw input. The two matrices can be concatenated along the first axis, resulting in a combined matrix (157 × 192). The combined matrix is then processed by a set of recurrent bi-directional layers (LSTM or GRU) followed by a temporally distributed dense layer with sigmoid activation. The resulting output vector (157 × 1) has values ranging from 0 (no cough) to 1 (cough).

To transform the final output into discrete cough timestamps, a peak-finding function is applied. This peak-finding function contains multiple tunable parameters and creates timestamps for all peaks exceeding a specified threshold. The final cough detection algorithm is based on an ensemble model of 16 independently trained models. The ensemble model combines the output vectors from the individual models by averaging them before applying the peak-finding function.

Model training and testing were conducted on fully independent datasets. Each sub-model is evaluated on a 4-fold cross-validation dataset. Additionally, the final ensemble model is divided into four sub-ensembles, with each sub-ensemble evaluated on its unseen data. Finally, an additional test is conducted to confirm the model’s performance on audio from the target device and to determine the final decision threshold when using the model in an ensemble (as optimal thresholds can vary across audio sources). The final ensemble is evaluated on a new, independent holdout test set to verify that the combined model outperforms its individual components. Each trained model is evaluated on a test set composed exclusively of data collected using SIVA’s proprietary device as part of usability testing, as required by the ISO 13485 standard. Evaluation is performed on complete 24-hour subject recordings (n = 11), without data augmentation or segment extraction. Overlapping predictions is averaged to produce the final output, which is then compared to the ground truth to compute performance metrics. After this model development, this cough detection algorithm was fixed for the final clinical validation.

## Confidence score calculation

To automatically flag patients for whom the cough detection algorithm may perform worse than expected, a confidence score is determined. Quantifying model prediction uncertainty based on the disagreement of the individual models in an ensemble is a standard approach in machine learning (Guo et al. 2017). For our purpose, we combined the standard deviation between the different individual model output vectors with the mean output vector, resulting in a cough event probability with associated uncertainty. A low standard deviation at the time of a detected potential cough event indicates high agreement between the individual models in the ensemble. Combined with a high mean output of the ensemble (i.e., a high probability for the presence of a cough event), this results in a high confidence value being assigned to the cough event. Conversely, a high standard deviation or a low ensemble output would contribute to a lower confidence value. Confidence values range from 0 (no confidence) to 1 (maximal confidence). These values were combined into a confidence score for a full day of a patient’s cough history by averaging the confidence values of all cough events detected on that day. A low daily confidence score, therefore, signifies a high level of disagreement between the individual cough detection models within the ensemble model on a given day.

This may indicate prolonged periods of cough detection uncertainty due to environmental influences on that day or a patient whose cough characteristics are “controversial” for the individual model of the ensemble model. In either case, it should be questioned whether the algorithm can maintain its expected detection performance. To support the analysis of the relationship between confidence score and algorithm performance, we heuristically defined a daily confidence score of less than 0.7 as “low confidence”, a score greater or equal 0.7 and less than 0.8 as “medium confidence”, a score greater or equal 0.8 and less than 0.9 as “high confidence”, and a score greater or equal 0.9 as “very high confidence”.

### Clinical validation study design

The final performance of the fixed cough detection algorithm was determined as part of a clinical validation study. The clinical validation study was designed as an interventional device study with a prospective, non-controlled, open-label design (NCT05689307). IRB approval was obtained from Advarra on November 2, 2022. The study was conducted at the Bellingham Asthma and Allergy Clinic in Washington, USA, in accordance with the Declaration of Helsinki, and all participants provided written informed consent. Patients eligible for the study included those with a cough lasting at least 12 months and persisting over the previous 8 weeks, with a diagnosis or suspicion of refractory chronic cough. All patients were recruited between March 2023 and November 2024 at a single site and were requested to use the SIVA System as part of the study. At visit 1, participants received the SIVA wearable (with charger), a smartphone pre-installed with the SIVA App, and a portable Wi-Fi device, and were instructed to use the system for 14 days to monitor their cough. They were asked to rate their current cough severity using a Visual Analog Scale (Cough Severity VAS) (Martin Nguyen et al. 2021) and assess their quality of life using the Leicester Cough Questionnaire (Birring et al. 2003), both at the beginning and end of the 14-day measurement period, with the final assessment conducted via phone interview. At the conclusion of the study, all equipment was returned to the clinic either in person or by mail. The first 24 hours of the audio data collected by the SIVA device in the beginning of the study were stored in accordance with the patient’s consent to enable validation of the cough detection algorithm. For the remainder of the study period, only the timestamps of the cough events were stored as data. The sample size was estimated using a bootstrap resampling method based on data from a prior clinical validation study in chronic cough patients (Kuhn et al. 2023) to achieve the expected device performance of greater than 82% Sensitivity and greater than 82% Positive Predictive Value with a 95% confidence interval across patients.

To evaluate the cough detection performance, each cough explosion was annotated by a trained human labeller at the exact start of the cough explosion phase (considered a true cough). This was determined by listening to the collected audio data supported by a visual inspection of a Mel Spectrogram. If a cough contained multiple explosions, the start of each explosion was labelled. Two independent human labellers labelled the data. In case of a disagreement, a decision from a third labeller was used based on a majority vote. The human-labelled cough timestamps were then compared to the output from the cough detection algorithm to determine the performance metrics of sensitivity (recall), positive predictive value (precision), F1 score, and false positive rate for each individual patient. Specificity was not considered a suitable metric for evaluating the performance of a cough detection algorithm due to the existence of large amounts of non-cough events, which would lead to misleadingly high specificity. Thus, precision was used instead as a metric that relies on true positive and false positive events.

The performance metrics were calculated as follows:

- Sensitivity (Recall) = (true positives) / (true positives + false negatives)
- Positive Predictive Value (Precision) = (true positives) / (true positives + false positives)
- F1 Score = 2 * [Recall * Precision / (Recall + Precision)]
- False Positive Rate = false positives / Cough Monitoring Time (cough per hour)

### Simulation to quantify the impact of monitoring time and algorithm performance on capturing true cough burden

To evaluate the effect of monitoring duration on cough burden estimation, we conducted a bootstrap-based simulation using up to 14 days of continuous cough monitoring data collected during the clinical validation study. For each patient, daily cough frequency (coughs/hour) was calculated over the available 24-hour recordings. The patient-specific reference value, referred to as the *true cough burden*, was defined as the mean daily cough frequency across the full monitoring period. Three patients with low-confidence algorithm performance were excluded from the simulation.

For each number of measurement days × ∈ [1,14], we sampled 10,000 random subsets of × days with replacement from each patient’s daily cough frequency data. We applied a measurement error model reflecting the patient’s observed algorithm performance on day one. Each sample’s average cough frequency was compared to the true cough burden to compute an absolute error; the mean of these absolute errors across samples gave the patient’s MAE, which was then normalized by their true cough burden to yield a *relative MAE*. These values were then averaged across all patients to obtain the **Average Relative MAE** for each X. As a baseline, the entire simulation was repeated without applying measurement error, simulating a hypothetical gold standard with perfect cough measurement.

## Code availability

The cough detection algorithm code used in the study is proprietary and is not publicly available.

## Data availability

Data are not publicly available because they contain sensitive patient information. Individual, de-identified participant cough data gathered in the clinical trial reported in this article may be made available to qualified researchers upon reasonable request. Data requestors will be required to sign a data-sharing agreement prior to access.

## Acknowledgements

The authors thank all study participants for their time and commitment to this research.

We are especially grateful to Jill S. Elkayam, Lead Clinical Research Department Coordinator at Bellingham Asthma and Allergy, and Samantha L. Jackson, Lead Clinical Coordinator on this project, for their outstanding efforts in patient recruitment and engagement throughout the study.

We also acknowledge the valuable contributions of the cough labelers for their meticulous work in the manual annotation of cough events, which was critical to the algorithm’s clinical validation. Finally, we thank Rael Kalda for assistance with data entry and Daniel Gisler for his support in data analysis.

## References

Abdulqawi, Rayid, Rachel Dockry, Kimberley Holt, et al. 2015. “P2X3 Receptor Antagonist (AF-219) in Refractory Chronic Cough: A Randomised, Double-Blind, Placebo-Controlled Phase 2 Study.” Lancet (London, England) 385 (9974): 1198–205. 10.1016/S0140-6736(14)61255-1.

Birring, S. S., T. Fleming, S. Matos, A. A. Raj, D. H. Evans, and I. D. Pavord. 2008. “The Leicester Cough Monitor: Preliminary Validation of an Automated Cough Detection System in Chronic Cough.” The European Respiratory Journal 31 (5): 1013–18. 10.1183/09031936.00057407.

Birring, S. S., B. Prudon, A. J. Carr, S. J. Singh, M. D. L. Morgan, and I. D. Pavord. 2003. “Development of a Symptom Specific Health Status Measure for Patients with Chronic Cough: Leicester Cough Questionnaire (LCQ).” Thorax 58 (4): 339–43. 10.1136/thorax.58.4.339.

Chaccour, Carlos, Isabel Sánchez-Olivieri, Sarah Siegel, et al. 2025. “Validation and Accuracy of the Hyfe Cough Monitoring System: A Multicenter Clinical Study.” Scientific Reports 15 (1): 880. 10.1038/s41598-025-85341-3.

Chung, Kian Fan, Carlos Chaccour, Lola Jover, et al. 2024. “Longitudinal Cough Frequency Monitoring in Persistent Coughers: Daily Variability and Predictability.” Lung 202 (5): 561–68. 10.1007/s00408-024-00734-x.

Dicpinigaitis, Peter V., Alyn H. Morice, Jaclyn A. Smith, et al. 2023. “Efficacy and Safety of Eliapixant in Refractory Chronic Cough: The Randomized, Placebo-Controlled Phase 2b PAGANINI Study.” Lung 201 (3): 255–66. 10.1007/s00408-023-00621-x.

Gemmeke, Jort F., Daniel P. W. Ellis, Dylan Freedman, et al. 2017. “Audio Set: An Ontology and Human-Labeled Dataset for Audio Events.” 2017 IEEE International Conference on Acoustics, Speech and Signal Processing (ICASSP), March, 776–80. 10.1109/ICASSP.2017.7952261.

Gross, Volker, Patrick Fischer, Andreas Weissflog, Olaf Hildebrandt, Ulrich Koehler, and Keywan Sohrabi. 2019. “Validation of the LEOSound Cough Detection Algorithm.” Preprint, Research Square, August 3. 10.21203/rs.2.12389/v1.

Guo, Chuan, Geoff Pleiss, Yu Sun, and Kilian Q. Weinberger. 2017. “On Calibration of Modern Neural Networks.” In Proceedings of the 34th International Conference on Machine Learning, edited by Doina Precup and Yee Whye Teh, vol. 70. Proceedings of Machine Learning Research. PMLR. https://proceedings.mlr.press/v70/guo17a.html.

Kuhn, Manuel, Elif Nalbant, Dario Kohlbrenner, et al. 2023. “Validation of a Small Cough Detector.” ERJ Open Research 9 (1): 00279–02022. 10.1183/23120541.00279-2022.

Martin Nguyen, Allison, Elizabeth D. Bacci, Margaret Vernon, et al. 2021. “Validation of a Visual Analog Scale for Assessing Cough Severity in Patients with Chronic Cough.” Therapeutic Advances in Respiratory Disease 15: 17534666211049743. 10.1177/17534666211049743.

McGarvey, Lorcan, Mandel Sher, Yury Grigorievich Shvarts, et al. 2023. “The Efficacy and Safety of Gefapixant in a Phase 3b Trial of Patients with Recent-Onset Chronic Cough.” Lung 201 (2): 111–18. 10.1007/s00408-023-00606-w.

Mines, Daniel, Elizabeth Bacci, Allison M. Nguyen, Shannon Shaffer, Jaclyn Smith, and Margaret Vernon. 2019. “Assessment of Inter- and Intra-Rater Reliability of Objective Cough Frequency in Patients with Chronic Cough.” Monitoring Airway Disease. European Respiratory Journal 54 (Suppl 63). 10.1183/13993003.congress-2019.PA4342.

Mohammadi, Sadegh Seyed, Steffen Vogler, Matthias Lenga, Daniel Rechsteiner, Mitja Alge, and Alexander Duschau-Wicke. 2024. Body action detection, identification and/or characterization using a machine learning model. European Union Patent EP4401628A1, filed September 8, 2022, and issued July 24, 2024. https://patents.google.com/patent/EP4401628A1/en.

Morice, Alyn H., Mitja Alge, Laura Kuett, Simon Hart, Alan Rigby, and David Elkayam. 2025. “Cough Frequency Has a High Daily Variation in Patients with Chronic Cough.” ERJ Open Research 11 (1): 00670–02024. 10.1183/23120541.00670-2024.

Morice, Alyn H., G. A. Fontana, M. G. Belvisi, et al. 2007. “ERS Guidelines on the Assessment of Cough.” The European Respiratory Journal 29 (6): 1256–76. 10.1183/09031936.00101006.

Morice, Alyn H., Eva Millqvist, Kristina Bieksiene, et al. 2020. “ERS Guidelines on the Diagnosis and Treatment of Chronic Cough in Adults and Children.” The European Respiratory Journal 55 (1): 1901136. 10.1183/13993003.01136-2019.

O’Hara, James, Holly Fisher, Louise Hayes, and Janet Wilson. 2022. “‘Persistent Throat Symptoms’ versus ‘Laryngopharyngeal Reflux’: A Cross-Sectional Study Refining the Clinical Condition.” BMJ Open Gastroenterology 9 (1): e000850. 10.1136/bmjgast-2021-000850.

Orlandic, Lara, Tomas Teijeiro, and David Atienza. 2023. “A Semi-Supervised Algorithm for Improving the Consistency of Crowdsourced Datasets: The COVID-19 Case Study on Respiratory Disorder Classification.” Computer Methods and Programs in Biomedicine 241 (November): 107743. 10.1016/j.cmpb.2023.107743.

Smith, Jacky, Kimberley Holt, and Kevin Mcguinness. 2021. “RADAR Database of Anonymised Acoustic Cough Recordings.” Manchester University NHS Foundation Trust.

Song, Woo-Jung, Yoon-Seok Chang, Shoaib Faruqi, et al. 2015. “The Global Epidemiology of Chronic Cough in Adults: A Systematic Review and Meta-Analysis.” The European Respiratory Journal 45 (5): 1479–81. 10.1183/09031936.00218714.

